# Suicide mortality in eastern and western Germany: construction of consistent time series 1952 to 2022

**DOI:** 10.1101/2025.10.14.25337714

**Authors:** Enno Nowossadeck, Claudia Hövener, Niels Michalski

## Abstract

Suicide mortality is a significant public health problem. Retrospective analyses can be helpful in better understanding temporal trend of suicide mortality and thus derive clues about causal and influencing factors for the course. For this paper, we compiled suicide mortality rates for eastern and western Germany from 1952 to 2022 from various data sources, stratified by sex. The period encompasses events that could substantially alter the statistically recorded rates, such as the changes in versions of the International Classification of Diseases (ICD) in 1978 and 1997, but also the integration of the statistical system of the German Democratic Republic (GDR) into that of the Federal Republic of Germany (FRG) in 1990/91. These could cause methodologically induced breaks in the compiled time series. Using Joinpoint-Jump models, we test whether at these defined points a jump, i.e., an abrupt change in the level of the time series, can be statistically detected or rejected. The empirical results show a high dynamism of suicide mortality in both parts of Germany, with very frequent increases and decreases of suicide mortality rates. However, the analyses show no evidence of methodologically induced structural breaks resulting from ICD version changes or from transferring of the GDR statistical system to that of the FRG. The analysis results support the conclusion that the compiled long time series for eastern and western German women and men are consistent. Therefore, future trend changes or shifts identified in analyses should not be attributed to methodological artefacts.

## 1. Introduction

Suicides are a major public health problem. Analyses of the World Mental Health (EU-WMH) survey in Germany revealed a lifetime prevalence of suicide ideation of 9.7% among adults, 2.2% for suicide plans and 1.7% for suicide attempts (Boyd et al., 2015; Nock et al., 2008). A recent study found an average lifetime prevalence of suicidal thoughts of 7.8% for the period 2003-2020 (Otten et al., 2024), with no significant differences measured between women and men. Since 1990, almost 360,000 suicide deaths have been registered in Germany – approximately 80% in western Germany, and around 20% in eastern Germany (Statistisches Bundesamt (Hrsg.), 2024). Small-scale differences and temporal dynamics exist within the two regions (Schelhase, 2022). At the time of German reunification in 1990, suicide mortality in eastern Germany was considerably higher than in western Germany and in many other European countries (Casper et al., 1990; Dinkel & Görtler, 1994; Wiesner, 2004). In the following years, suicide mortality declined in both parts of Germany, and this continued until around 2007 (Hegerl et al., 2013). In view of this temporal dynamic and unclear causal mechanisms, a retrospective analysis of suicide mortality in Germany is of great interest. However, the availability of consistent time-series data is a mandatory prerequisite for such an analysis. The existence of methodologically induced structural breaks in the periods to be analysed would represent a problem to be taken into account in the analysis. One example of this is the version changes to the International Classification of Diseases (ICD). Each revision can cause breaks in the comparability of the data (Hetzel, 1997), and this can have a negative impact on the consistency of the time series of the analysed causes of death.

Another factor that can impact on the consistency of the time series is the transfer of the official statistics system of the German Democratic Republic (GDR) to that of the Federal Republic of Germany (FRG) in the course of reunification in 1990. Starting in October 1990, the statistical system in eastern Germany was structurally and conceptually reorganized and standardized for the whole of Germany. Thus, data sets on the cause-of-death statistics for eastern and western Germany, collected according to standardized regulations and guidelines, have been available for analysis since 1991 (Sommer & Voit, 1998). As pertinent analyses have shown, there were also different coding practices and customs in the GDR and the FRG despite their using the same ICD versions (Wengler et al., 2019).

This shows that methodological consistency of the statistical time-series data required for trend analyses – including the reunification period – should not be assumed *a priori*. The problems of integrating GDR statistics into those of the FRG have been described for some statistical indicators, for example for national accounts (Heske, 2009).

To assess the usability of the data for analysing temporal trends, this paper pursues the following objectives:

a. to merge time series for eastern and western Germany for the period from 1952 to 2022 from various data sources, and
b. to check for any methodologically induced structural breaks and to ensure that no such breaks in time series exist in future analyses.
c. 2. Data and methods

### 2.1. Data

For our analysis, we used data from the national cause-of-death statistics of the GDR and the FRG. The compilation and coding of suicide deaths in the cause-of-death statistics is standardized internationally using the ICD. This coding system, developed by the WHO, has been the basis of the statistical systems of the GDR and the FRG in the latest versions of cause-of-death documentation since the 1950s.

The ‘European Shortlist for Causes of Death’ published by Eurostat is available for historical comparisons. It combines the deaths from groups of diseases and external causes in the ICD versions 8, 9 and 10 (Eurostat, 2012). In the case of suicide deaths, the category is called ‘Suicide and intentional self-harm**’**.

When the official statistics system of the GDR was transferred to that of the FRG in 1990/1991, data from the cause-of-death statistics for previous years were also transferred. These can be retrieved retrospectively up to 1980 from the online health-reporting database (https://www.gbe-bund.de/gbe). For the analyses, we compiled further data for the period from 1952 to 1979 from various sources. For the FRG, data is available from the 1952-1979 Statistical Yearbooks (Statistisches Bundesamt (Hrsg.), 1952–1979). Data on suicide deaths were collected annually in the GDR; however, they were systematically withheld from public release starting in 1972 to 1989 (Casper et al., 1990; Hoffmeister et al., 1990; von den Driesch, 2021) (cf. Table 1).

**Table 1.** Compilation of sources for eastern and western Germany in different time periods.

| Period | Eastern Germany | Western Germany | Period |
| --- | --- | --- | --- |
| 1952-1979 | GESIS Archive Research Data<br>(von den Driesch, 2020) | Statistical yearbooks<br>(Statistisches Bundesamt<br>(Hrsg.), 1952–1979) | 1952-1979 |
| 1980-2022 | Online database for health reporting<br>(Statistisches Bundesamt<br>(Hrsg.), 2024) | Online database for health reporting<br>(Statistisches Bundesamt<br>(Hrsg.), 2024) | 1980-2022 |

Source: own compilation

GDR data for the period 1952-1979 were included in the present analyses. It therefore seems appropriate to examine whether the change of data source in 1979/1980 introduced a methodologically induced structural break, since the possibility of incomplete datasets cannot be excluded.

A methodologically induced structural break is defined as methodological changes in the compilation, coding and/or aggregation of data that change the previous trend of the time series or its level. It is irrelevant whether the new level is higher or lower than the former one. Such discontinuities or ‘jumps’ in the data can potentially lead to misleading interpretations if they are not taken into account (Chen et al., 2020).

There may be methodologically induced structural breaks in the suicide mortality data for several year pairs. Table 2 shows these year pairs, keeping eastern and western Germany separate.^1^

**Table 2.** Timing of possible methodologically induced structural breaks.

| East |  | West |  |
| --- | --- | --- | --- |
| Year | Event | Year | Event |
| 1977/78 | Version switch from ICD8 to ICD9 | 1977/78 | Version switch from ICD8 to ICD9 |
| 1979/80 | Change in the data source |  |  |
| 1990/91 | Reorganization of official statistics |  |  |
| 1997/98 | Version switch from ICD9 to ICD10 | 1997/98 | Version switch from ICD9 to ICD10 |

Source: own compilation

The analyses included the crude rates of suicide mortality (deaths per 100,000 of the population) for eastern and western Germany, stratified by gender.

### 2.2. Hypotheses

This article examines whether there are methodologically induced structural breaks. Our overarching hypothesis H is therefore as follows: there are no methodologically induced structural breaks in the gender-specific time series for eastern and western Germany at the points in time mentioned in Table 2. The following hypotheses are tested for the relevant groups in separate statistical analyses:

**Hypothesis H1a.** Our first hypothesis is that there is no methodologically induced structural break in 1977/78 due to the respective ICD version change.
**Hypothesis H1b.** Analogous to H1a, we assume that there is no methodologically induced structural break in 1997/98.
**Hypothesis H2.** Our second hypothesis assumes that there is no methodologically induced structural break in the GDR time-series data for 1979/80 as a result of the change in data access.
**Hypothesis H3.** The third hypothesis is that there is no methodologically induced structural break in the time-series data for 1990/91 as a result of the reorganization of official statistics in eastern Germany.

### 2.3. Methods

The suicide rates were retrieved separately for women and men from the above-mentioned databases and collated for FRG/GDR and west/east respectively. This resulted in four time series: women in western and in eastern Germany and men, also in western and eastern Germany.

Joinpoint-jump model calculations were carried out for the compiled time series. The joinpoint analysis is based on the idea of visualizing trends in time series through a sequence of connected linear sections. The optimum position and number of joinpoints (or linear segments) is determined by model comparisons using the Bayesian Information Criterion 3 (Kim et al., 2023). The procedure tests all possible joinpoints positions within the data, and the model with the best fit.

In addition, joinpoint-jump models estimate whether there is a sudden jump within one of the linear trend segments identified by the joinpoint regression, resulting, for example, from a change in health-statistics coding (Chen et al., 2020). Models were calculated with jumps in the year pairs specified in Table 2. So-called ‘comparability ratios’ (CRs) were calculated to statistically verify whether the jumps indicate a break in the respective time series. The CRs show the ratio of the rates estimated by the linear trend before and after the jump (Chen et al., 2020). If the CR equals 1, the rates are unaffected by the change in method (such as an ICD version change, etc.). In this case, there is no methodologically induced structural break. If the CR is larger than 1, the rates after the method change are higher than before, and vice versa: if CR is less than 1, the rates after the method change are lower than before. The joinpoint-jump models therefore only make it possible to identify a methodologically induced structural break. However, a correct inference requires that the point in time at which the joinpoint is assumed should not be close to or at the end of a segmented trend, as there is then a risk of confusion between joinpoint and jump. It should be noted that a jump can only be identified within a time interval between two joinpoints. However, as the number of joinpoints changes across models, the time periods defined by the joinpoints can also change. The models are only comparable with each other with regard to the jumps if the time periods in which the jump is assumed do not differ significantly between models.

In conducting a content analysis, it is necessary to test the hypothesis that the data are free of methodologically induced structural breaks. If there are no such structural breaks, we can assume that the compiled time series are consistent. Results with p-values larger than 0.05 and less than 0.20 thus remain inconclusive.

The maximum number of seven joinpoints follows the recommendation to select seven joinpoints as the default setting for a time-series length of at least 37 points in time (National Cancer Institute, 2023). 71 points in time are available in each of the time series we analysed. In the results tables, the results for two to seven joinpoints are shown for all models to gain a deeper understanding; in the figures and in the discussion, we use the results of the models with seven joinpoints.

We use this procedure to check whether there are significant values for the CRs, i.e. significant jumps, for the specified year pairs. A methodologically induced structural break is assumed if the following criteria apply:

a. The prerequisite for testing is that the time point under examination for the time series falls approximately within the same trend segment.
b. All jumps move in the same direction for one point in time that is to be tested, i.e. all relevant comparability ratios of a point in time are either greater than 1 (increase) or less than 1 (decrease).
c. The comparability ratios are significant for the respective point in time in all populations studied (women/men x eastern/western Germany).
d. The hypothesis that there is <u>no</u> significant jump cannot be rejected (test beta error).

## 3. Results

Combining the data from the different data sources makes time series of crude suicide-mortality rates available for the period from 1952 to the present. These are shown graphically in Figure 1.

**Figure 1:**
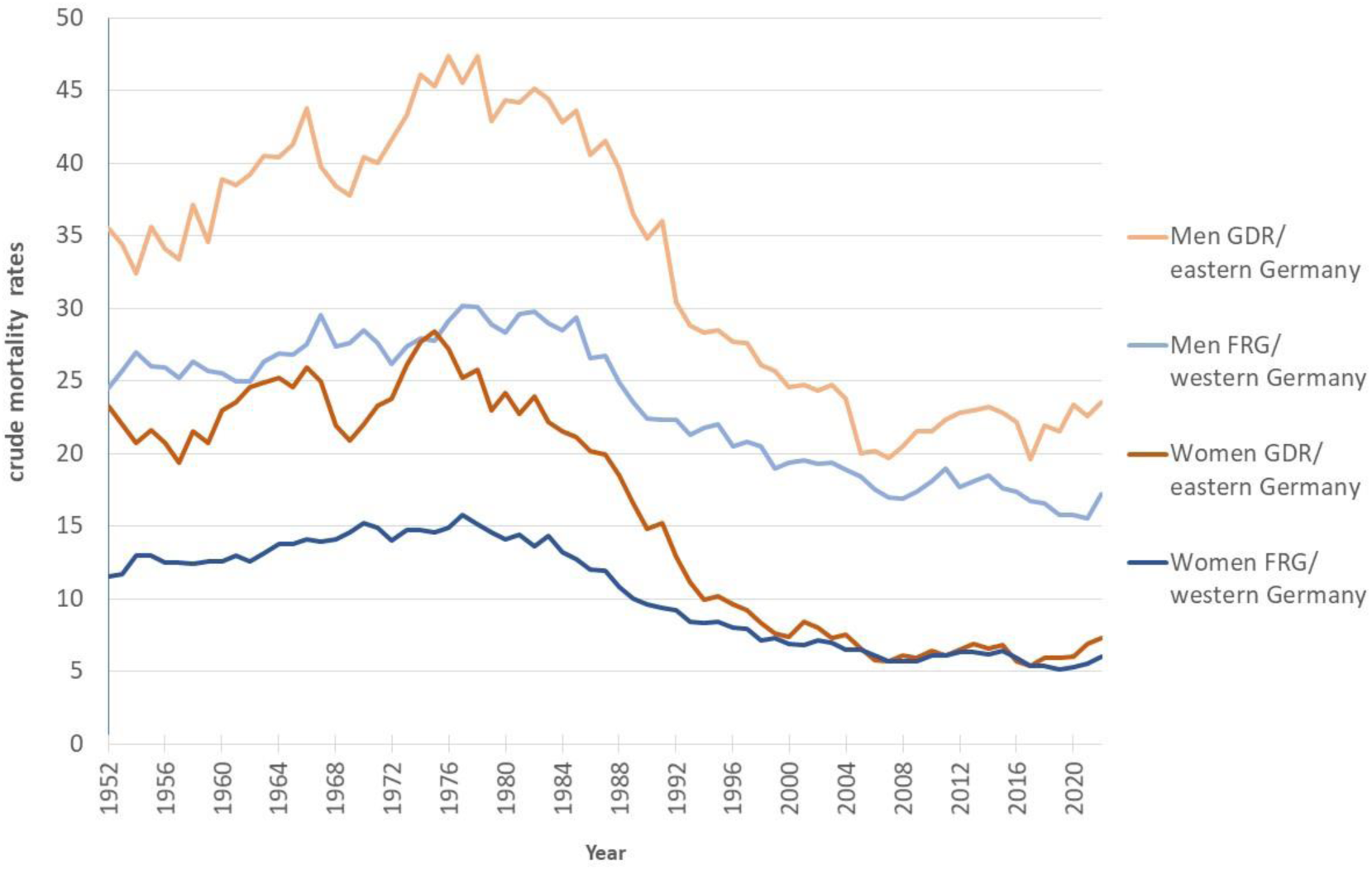
Suicide mortality in eastern and western Germany 1952-2022: presentation of the compiled time series, suicide deaths per 100,000 of the population, women and men. Source: own calculations Data sources: (Statistisches Bundesamt (ed.), 1952-1979, 2024; von den Driesch, 2020)

Suicide mortality in eastern Germany is higher than in western Germany, for both women and men. However, since the turn of the millennium, there have been no – or only slight – differences between women in the two parts of the country. Over the entire period, the suicide mortality rate is higher among men than among women.

Ignoring annual fluctuations, the temporal development can be divided into three phases with distinct quantitative characteristics:

- Phase I: up until the first half of the 1980s, suicide mortality rates rose among both women and men.
- Phase II: this was followed by a decline in suicide mortality rates in both parts of Germany and among both women and men, which continued into the first half of the 2000s.
- Phase III: a plateau phase began around the mid-2000s.

The decline during Phase II was greater in eastern Germany than in the western part, for both women and men.

Tables 3 to 6 contain the results of the joinpoint-jump models with two to seven joinpoints for each of the year pairs shown in Table 2. As a rule, the models with two joinpoints reflect the three phases (with the exception of men in western Germany), thus indicating the phase in which the jump is empirically located. The joinpoint-jump models are discussed separately for the year pairs below.

**Table 3.** Linear segment of the JP model in which the jump of the 1977/78 year pair falls, and the corresponding CR.

| Number<br>of<br>joinpoints | East |  |  |  |  |  | West |  |  |  |  |  |
| --- | --- | --- | --- | --- | --- | --- | --- | --- | --- | --- | --- | --- |
|  | Women |  |  | Men |  |  | Women |  |  | Men |  |  |
|  | Trend<br>segment | CR | p-value<br>of the CR | Trend<br>segment | CR | p-value<br>of the<br>CR | Trend<br>segment | CR | p-value<br>of the<br>CR | Trend<br>segment | CR | p-value<br>of the CR |
| 2 | 1952-1982 | 0.914 | 0.099 | 1952-1982 | 0.950 | 0.130 | 1952-1981 | 0.918 | 0.009 | 1952-1985 | 0.994 | 0.786 |
| 3 | 1952-1983 | 0.923 | 0.093 | 1952-1983 | 0.950 | 0.097 | 1952-1981 | 0.933 | 0.022 | 1952-1985 | 0.993 | 0.730 |
| 4 | 1976-1987 | 1.023 | 0.805 | 1952-1982 | 0.951 | 0.114 | 1952-1978 | 0.982 | 0.450 | 1952-1985 | 0.994 | 0.755 |
| 5 | 1976-1987 | 1.023 | 0.796 | 1952-1983 | 0.970 | 0.073 | 1952-1981 | 0.932 | 0.003 | 1952-1985 | 0.994 | 0.738 |
| 6 | 1976-1987 | 1.023 | 0.780 | 1975-1987 | 1.013 | 0.774 | 1977-1983 | 0.972 | 0.386 | 1952-1985 | 0.993 | 0.692 |
| 7 | 1975-1988 | 1.014 | 0.828 | 1976-1987 | 1.031 | 0.546 | 1977-1983 | 0.978 | 0.484 | 1967-1985 | 1.023 | 0.406 |

### 3.1. 1977/78 year pair

The 1977/78 year pair lies in the final temporal section of the first phase (see Fig. 2). Depending on the number of permitted joinpoints, the joinpoint-jump models identify different trend segments into which the year pair falls (see Table 3).

**Figure 2.**
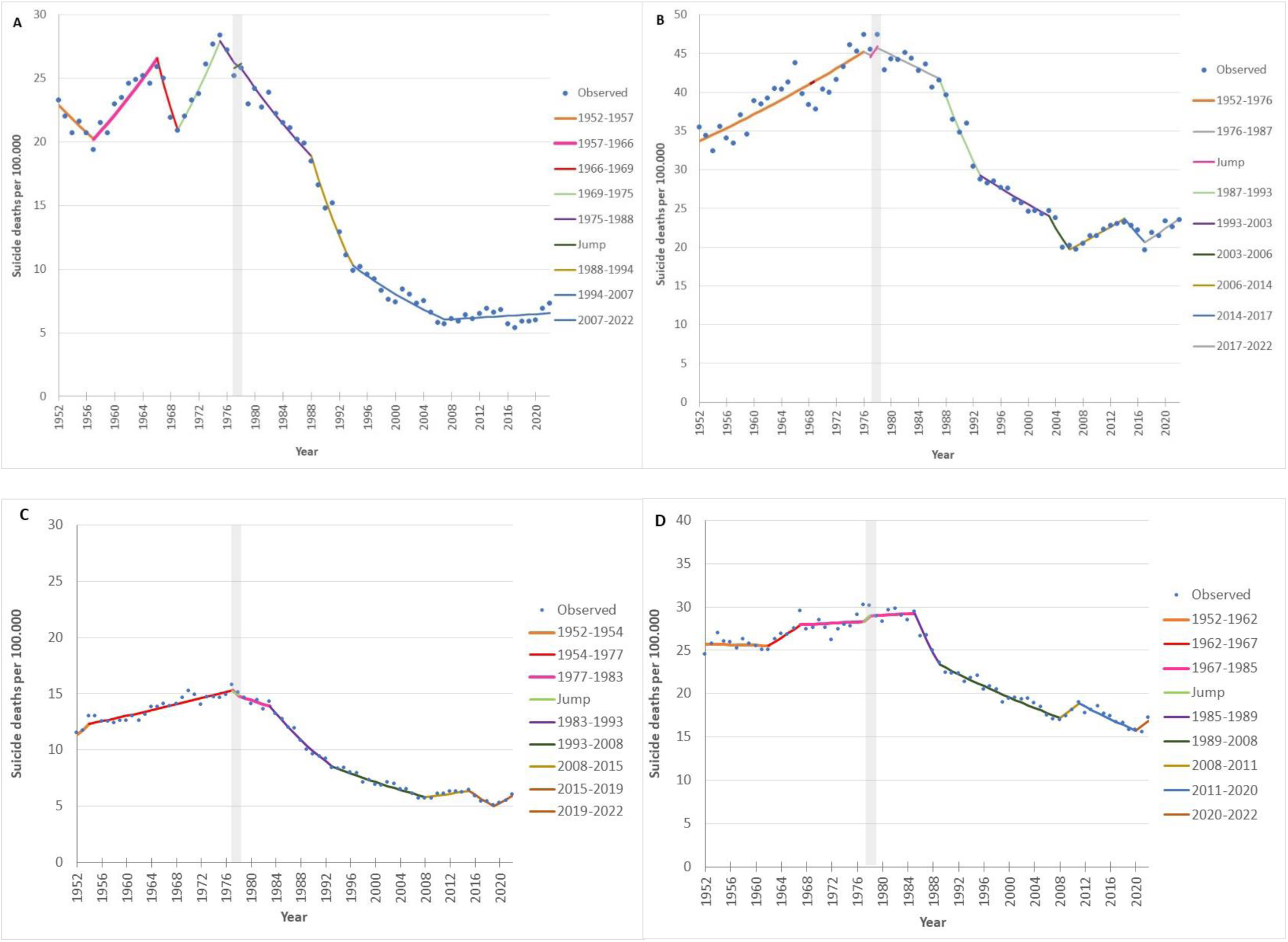
Results of the joinpoint-jump modellings, 1977/78. A) Women, eastern Germany. B) Men, eastern Germany. C) Women, western Germany. D) Men, western Germany Source: own calculations; model selection method: BIC3; maximum number of joinpoints: 7; presentation: models with 7 joinpoints Data sources: (Statistisches Bundesamt (ed.), 1952-1979, 2024; von den Driesch, 2020)

In models with a small number of joinpoints, the year pair falls into a trend segment from 1952 to 1981/85, which represents the whole of Phase I. Models with a higher number of joinpoints generate a trend segment that extends from the mid-1970s to the mid-1980s. The jumps are not significant, with two exceptions: the models with 2 and 5 joinpoints for women in the west.

Both trend segments end in the first half of the 1980s, i.e. in the period in which the first phase ends. The beginning varies depending on whether only one or several trend segments are estimated for the first phase. In models in which the trend segment covers the entire first phase, the jumps are not significantly different from 1. In some cases (women east: 2, 3 joinpoints, men east: 2-5 joinpoints, women west: 3 joinpoints), the hypothesis of the absence of jumps cannot be rejected (p-value >=0.05 and < 0.2).

Overall, there are hardly any indications of a methodologically induced structural break for the 1977/78 year pair.

### 3.2. 1979/80 year pair

The 1979/80 year pair falls within the final section of the first phase (Fig. 3). The models with a small number of joinpoints assign the year pair to the trend segment that covers the entire phase I, especially for men (2-5), for women only the model with 3 joinpoints. Several jumps are significant in this specification (see Table 4).

**Figure 3:**
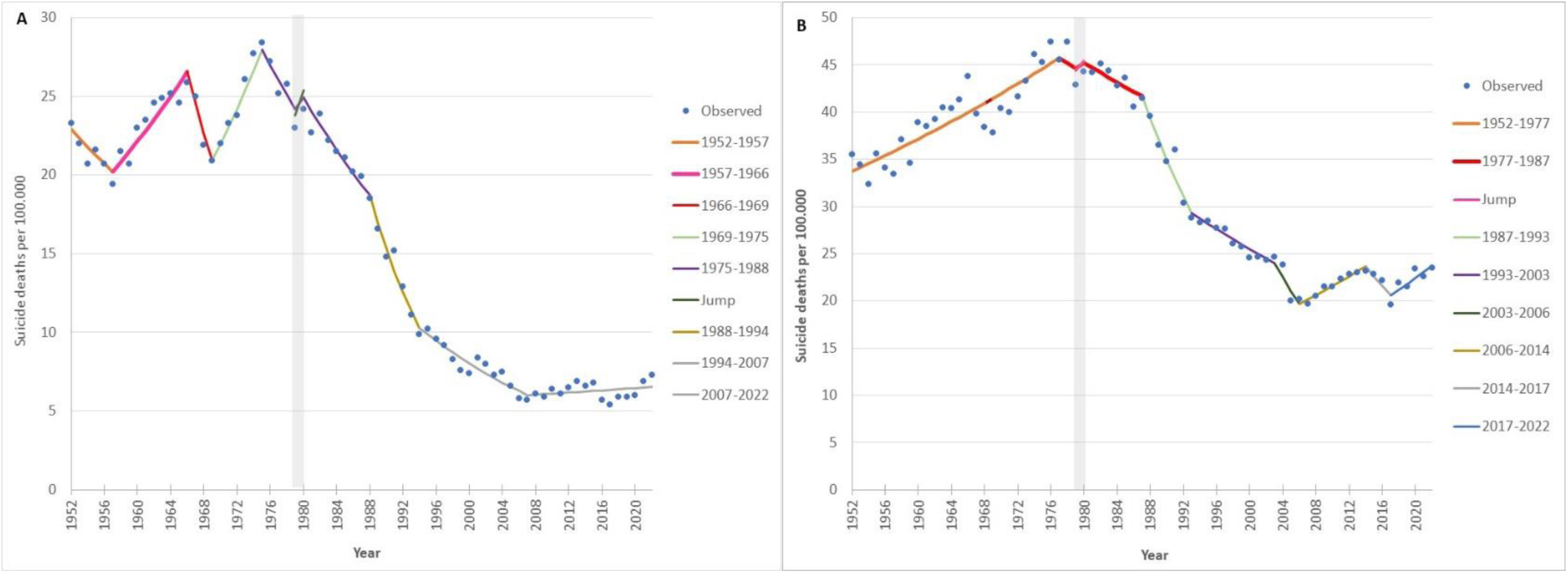
Results of the joinpoint-jump modellings, 1979/80. A) Women, eastern Germany. B) Men, eastern Germany. Source: own calculations; model selection method: BIC3; maximum number of joinpoints: 7; presentation: models with 7 joinpoints Data sources: (Statistisches Bundesamt (ed.), 1952-1979, 2024; von den Driesch, 2020)

**Table 4.** Linear segment of the JP model in which the jump of the 1979/80 year pair falls, and the corresponding CR.

| Number<br>of<br>joinpoints | East |  |  |  |  |  |
| --- | --- | --- | --- | --- | --- | --- |
|  | Women |  |  | Men |  |  |
|  | Trend<br>segment | CR | p-value<br>of the CR | Trend<br>segment | CR | p-value<br>of the<br>CR |
| 2 | 1978-2006 | 1.128 | 0.220 | 1952-1982 | 0.952 | 0.225 |
| 3 | 1952-1985 | 0.896 | 0.016 | 1952-1985 | 0.917 | 0.003 |
| 4 | 1976-1988 | 1.073 | 0.388 | 1952-1982 | 0.953 | 0.298 |
| 5 | 1976-1988 | 1.073 | 0.371 | 1952-1984 | 0.931 | 0.013 |
| 6 | 1976-1988 | 1.074 | 0.335 | 1976-1987 | 1.023 | 0.656 |
| 7 | 1975-1988 | 1.067 | 0.341 | 1977-1987 | 1.024 | 0.623 |

In other models, the year pair falls into trend segments that begin in 1975 and last until 1988 (women, models with 2, 4-6 joinpoints) or 1987 (men, models with 6 and 7 joinpoints). None of the jumps in these trend segments are significant.

The indications of a methodologically induced structural break are contradictory. On the one hand, three models show a significant jump that indicates a decline. On the other hand, all other models show no significant jumps, and the beta error test generates no evidence that discarding a jump would conceal relevant structural breaks (p-value > 0.2). Overall, there is no clear statistical evidence of significant jumps related to the 1979/80 year pair.

### 3.3. 1990/91 year pair

This year pair falls within the second phase, which is characterized by declining suicide mortality rates in all populations (Fig. 4). Two trend segments can be identified: a longer one extending from the early 1980s to the mid-2000s (women 2-3 joins; men 2-5 joins), and a much shorter one covering the period from 1988 to 1993/94 (women 4-7 joins, men 6-7 joins) (see Table 5).

**Figure 4:**
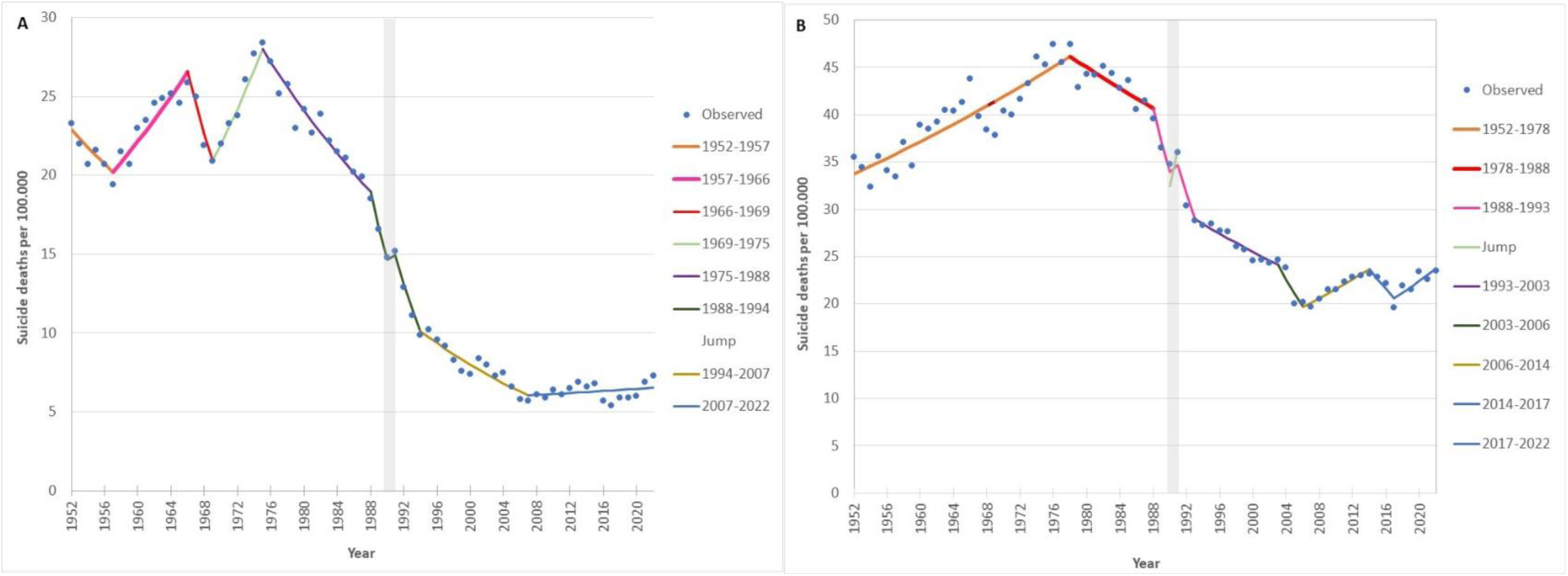
Results of the joinpoint-jump modellings, 1990/91. A) Women, eastern Germany. B) Men, eastern Germany. Source: own calculations; model selection method: BIC3; maximum number of joinpoints: 7; presentation: models with 7 joinpoints Data sources: (Statistisches Bundesamt (ed.), 1952-1979, 2024; von den Driesch, 2020)

**Table 5.** Linear segment of the JP model in which the jump of the 1990/91 year pair falls, and the corresponding CR.

| Number<br>of<br>joinpoints | **East** |  |  |  |  |  |
| --- | --- | --- | --- | --- | --- | --- |
|  | Women |  |  | Men |  |  |
|  | Trend<br>segment | CR | p-value<br>of the CR | Trend<br>segment | CR | p-value<br>of the<br>CR |
| 2 | 1980-2006 | 0.901 | 0.078 | 1981-2006 | 0.924 | 0.036 |
| 3 | 1980-2006 | 0.892 | 0.048 | 1983-2006 | 0.925 | 0.038 |
| 4 | 1988-1994 | 1.160 | 0.300 | 1981-2007 | 0.928 | 0.028 |
| 5 | 1988-1994 | 1.163 | 0.270 | 1983-2006 | 0.925 | 0.026 |
| 6 | 1988-1994 | 1.158 | 0.252 | 1988-1993 | 1.110 | 0.298 |
| 7 | 1988-1994 | 1.160 | 0.213 | 1988-1993 | 1.121 | 0.229 |

The jumps for the trend segments that begin in 1980/81 and are associated with a decline in suicide mortality rates are particularly significant.

In the case of shorter trend segments (from 1987/88), no significant jumps are observed. Similarly, there are no indications of a beta error, i.e. the erroneous rejection, based on the test statistics, of a jump that actually exists.

The evidence of a methodologically induced structural break are therefore somewhat contradictory. However, an examination of the graphs suggests that solutions with a higher number of joinpoints are preferable because they identify shorter trend segments in this area of the sharp decline in suicide mortality. The beginning and end of phase II, where the decline is not yet as strong or flattens out again, have a less distorting effect on the linear decline for all populations (cf. Fig. 4).

### 3.4. 1997/98 year pair

For 1997/98, the trend segments that include the jump vary considerably across the populations (Fig. 5). However, the majority of them indicate periods towards the end of the second phase. Trend segments that extend significantly beyond the second phase (1990-2022, 1994-2020, 1993-2022) are estimated for models with two or three joinpoints for men and women in the west. Here, the jumps are significant and indicate a decrease in suicide mortality (see Table 6).

**Figure 5:**
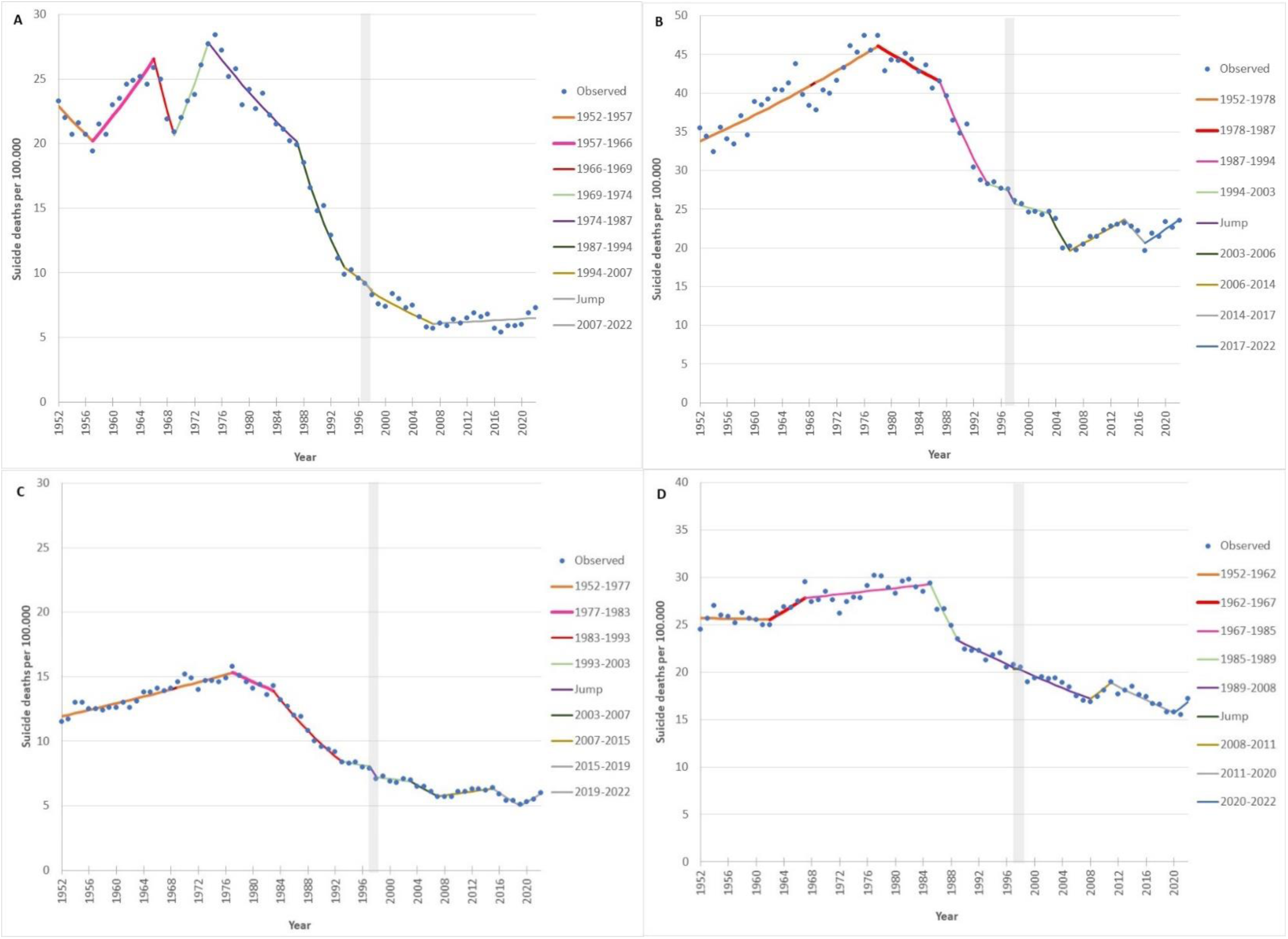
Results of the joinpoint-jump modellings, 1997/98. A) Women, eastern Germany. B) Men, eastern Germany. C) Women, western Germany. D) Men, western Germany Source: own calculations; model selection method: BIC3; maximum number of joinpoints: 7; presentation: models with 7 joinpoints Data sources: (Statistisches Bundesamt (ed.), 1952-1979, 2024; von den Driesch, 2020)

**Table 6.** Linear segment of the JP model in which the jump of the 1997/98 year pair falls, and the corresponding CR.

| Number<br>of<br>joinpoints | **East** |  |  |  |  |  | **West** |  |  |  |  |  |
| --- | --- | --- | --- | --- | --- | --- | --- | --- | --- | --- | --- | --- |
|  | Women |  |  | Men |  |  | Women |  |  | Men |  |  |
|  | Trend<br>segment | CR | p-value<br>of the CR | Trend<br>segment | CR | p-value<br>of the<br>CR | Trend<br>segment | CR | p-value<br>of the<br>CR | Trend<br>segment | CR | p-value<br>of the CR |
| 2 | 1980-2006 | 1.008 | 0.895 | 1982-2006 | 1.067 | 0.116 | 1994-2022 | 0.881 | 0.000 | 1990-2022 | 0.948 | 0.015 |
| 3 | 1982-1999 | 0.962 | 0.676 | 1985-2005 | 1.851 | 0.075 | 1994-2020 | 0.898 | 0.001 | 1990-2020 | 0.956 | 0.045 |
| 4 | 1994-2007 | 0.951 | 0.490 | 1982-2007 | 1.066 | 0.100 | 1978-2007 | 1.034 | 0.245 | 1989-2008 | 1.002 | 0.949 |
| 5 | 1994-2007 | 0.939 | 0.358 | 1985-2005 | 1.085 | 0.056 | 1993-2008 | 0.953 | 0.140 | 1989-2008 | 1.003 | 0.920 |
| 6 | 1994-2007 | 0.958 | 0.508 | 1993-2006 | 1.023 | 0.638 | 1993-2008 | 0.958 | 0.145 | 1990-2004 | 0.969 | 0.339 |
| 7 | 1994-2007 | 0.951 | 0.407 | 1994-2003 | 0.944 | 0.288 | 1993-2003 | 0.903 | 0.008 | 1989-2008 | 1.003 | 0.895 |

Source: own calculations; model selection method: BIC3; maximum number of joinpoints: 7; highlighted in grey: frequently identified trend segments. Data sources: (Statistisches Bundesamt (Hrsg.), 1952–1979, 2024; von den Driesch, 2020)

The models with a higher number of joinpoints (>3) often show trend segments around 1994-2007. A fall is estimated for women in these models (CR around 0.95). These falls are not significant either for women in eastern Germany (4-7 joinpoints) or in western Germany (5, 6 joinpoints). In the models for women in western Germany (5, 6 joinpoints), however, the hypothesis of the existence of a jump cannot be rejected either (p-value >=0.05 & < 0.2). In the model with 7 joinpoints, the fall in a slightly different sub-segment (1993-2003) is also estimated to be somewhat stronger and is significant.

For men in eastern Germany, such a decline is only evident in the model with 7 joinpoints, in this case without significance. For men in western Germany, the trend segments consistently begin somewhat earlier (1989/90) and no conspicuous CRs are estimated.

There are signs of a methodologically induced structural break for this year pair among women, but these are subject to statistical uncertainty.

Overall, the trend segments in which the jump was tested varied considerably with the number of joinpoints across the subpopulations and across the models. This also had an impact on the direction, strength and significance of the CRs. However, similar CRs were identified in the subpopulations for the same trend segments.

### 3.5. Hypothesis testing

#### Hypotheses 1a and 1b

Our initial hypotheses were that in the years 1977/78 and 1997/98 there would be no methodologically induced structural break in the time series data caused by the respective ICD version change. The result of the analyses was that there is hardly any evidence of a methodologically induced structural break associated with the two year pairs.

#### Hypothesis 2

This hypothesis stated that in 1979/80 there was no methodologically induced structural break in the time series data of the GDR caused by the change in data access. The results for the most frequently occurring trend segment (mid-1970s to the second half of the 1980s) show no significant jumps for this year pair (Tables 3-6). Taking into account the tests for beta errors, it can be concluded that no methodologically induced structural break can be detected for 1979/80. Hypothesis 2 is therefore not rejected.

#### Hypothesis 3

With hypothesis 3 we stated that the reorganization of official statistics in 1990/91 in eastern Germany did not lead to a methodologically induced structural break. We found no significant jumps in those trend segments that begin in 1988, but we do in those that begin in 1980. However, this year pair coincides with a phase of sharp decline in suicide mortality, which is clearly linear. Furthermore, the direction of the estimated CR is inconsistent, and the fall of the Berlin Wall in 1990 was a one-off event which in some cases was associated with increased suicide mortality. Although the evidence of a methodologically induced structural break are ambiguous here, overall, they can be described as insignificant.

## 4. Discussion

The first aim of the article was to merge time series from different data sources for eastern and western Germany for the period 1952 to 2022. These are now available.

- An initial analysis of suicide mortality trends shows the following: The development of suicide mortality can be divided into three phases for women and men in the two parts of Germany over the period analysed here. Phase I: increase in suicide mortality until the first half of the 1980s; Phase II: subsequent decline in suicide mortality until the mid-2000s; Phase III: plateau phase since the second half of the 2000s.
- Known differences in suicide mortality between eastern and western Germany (e.g (Dinkel, 1994; Hoffmeister et al., 1990; Wiesner & Haberland, 1996)) are confirmed.
- The three phases are similar in both parts of the country: although they are quantitatively different, their basic tendencies were the same.
- A downward trend has already been reported in previous studies, e.g. (Baumert et al., 2005; Casper et al., 1990; Hoffmeister et al., 1990), but only for parts of the period analysed here.
- The development of suicide mortality rates is characterized by very frequent increases and decreases and is thus highly dynamic in women and men across eastern and western Germany.
- There is no obvious evidence of methodologically induced structural breaks in the year pairs mentioned.

The second objective was to examine whether any methodologically induced structural breaks occurred in the analysed year pairs. The result of testing the three hypotheses was that there was no clear and reliable statistical evidence of a methodologically induced structural break in any of the year pairs analysed in this article. Nevertheless, there are some characteristics to be discussed relating to the respective year pairs.

### 4.1 1977/78 and 1997/98 year pairs: change in the ICD version

We found no reliable statistical evidence of a methodologically induced structural break in either of the year pairs as postulated in hypotheses 1a and 1b. This result is supported by the fact that the Eurostat European Short List contains a category ‘Suicide and deliberate self-harm’ across ICD versions (Eurostat, 2012). This shows that no methodologically induced break in the collection of these data is seen in the official statistics.

In the international literature analysing long time-series of suicide mortality, the topic of ICD version changes is only mentioned only sporadically, if at all (e.g. (Chishti et al., 2003; Dávila-Cervantes, 2022; Dyvesether et al., 2018)). In 2002, Bertolote & Fleischmann came to the conclusion that the category name and coding of suicide mortality are relatively stable across the different versions of the ICD (6-10) (Bertolote & Fleischmann, 2002)

We are not aware of any empirical reviews of these statements, which more or less explicitly regard the effects of ICD version changes on the analyses of suicide mortality as negligible. Overall, we therefore assume that the ICD version changes did not lead to methodologically induced structural breaks in suicide mortality in either part of Germany.

A recent study concluded that the GDR suicide statistics adequately depicted the development of suicide mortality (Grashoff, 2023). When processed by von den Driesch (von den Driesch, 2021) and the Federal Statistical Office (Statistisches Bundesamt (Hrsg.), 2024), all suicide mortality data that had kept secret in the GDR could apparently be incorporated without substantial gaps (Casper et al., 1990; Grashoff, 2006; Hoffmeister et al., 1990; von den Driesch, 2021).

### 4.2 1980/81 year pair: change in the data basis

Hypothesis 2 analysed the 1979/80 year pair in order to examine the possible effects of a change in the data basis. The year pair is close in time to the transition from phase I to phase II, during which the rise in suicide mortality ends and transitions into a decline. The application of the joinpoint-jump analysis assumes an uninterrupted general trend of the process analysed. Since this is not the case here, the results should be interpreted with caution. It should be noted that there was a profound change in the development of suicide mortality in both parts of Germany during this period, and this makes an unequivocal finding difficult. The hypothesis that the change of data source represents a methodologically induced structural break can therefore neither be confirmed nor definitively rejected.

### 4.3 1990/91 year pair: reorganization of official statistics

For the 1990/91 year pair, the temporal variation is relatively small in all groups (which may partly reflect the larger population size in the western federal states). The overall trend can therefore be regarded as stable. The CR is not significant for women and men in the new federal states or for men in the old federal states, but it is significant for women in the old federal states (at 7 joinpoints, cf. Table 6). At the outset, we determined that the CRs are significant for all time series (women, men, east, west) if there is a methodologically induced structural break present, e.g. as a result of the version change. For this year pair we conclude that the formulated hypothesis of a jump can be rejected. However, the beta error test shows that the associated hypotheses H1a and H1b (no methodologically induced structural break) are not fully supported by the test, because the p-values for the CRs are lower than 0.20 for women in the west. This means that the presence of a jump cannot be rejected, at least for this group. Given the strength of the decline in phase II, on the other hand, it can be ruled out that the data contain a particularly strong methodologically induced structural break.

What is empirically striking is that the suicide mortality rates in eastern Germany were higher in 1991 than in 1990 for both women and men, which appears significant against the background of a strong overall downward trend. In the literature, this temporary increase has been linked to the social changes in the course of German reunification (Casper et al., 1990; Schmidtke et al., 1999; Straub, 2000; Tasseit, 1993; Wiesner & Haberland, 1996). Wiesner and Haberland explicitly point out the impact on mental health of involuntary unemployment, which occurred on a large scale after 1990 (Wiesner & Haberland, 1996).

The interruption of the trend is therefore more likely to be due to the consequences of social change and the transformation process in eastern Germany.

### 4.4 Summarizing assessments

Table 7 summarizes the criteria for identifying a methodologically induced time-series break, as presented in the methods section, and the respective results. Column 1 lists the year pairs, and column 2 the number of subpopulations (east/west, female/male). This forms the basis for further tests of the respective trend segments (criterion 1). This is used to carry out further tests for the respective trend segment (criterion 1). Column 3 shows the two most frequently identified trend segments from Tables 3-6 (highlighted in grey). Column 4 shows how often each trend segment occurred in the respective year pair for all subpopulations. For example, for the 1977/78 year pair, the trend segment 1952 to 1981-85 occurs 15 times in all models (with 2-7 joinpoints each) and the trend segment 1975-77 to 1981-87 occurred 8 times. Thus, in 23 out of 24 models (6 models with different joinpoints x gender x east/west) two trend segments were identified. One model (model with 7 joinpoints, men, west: 1967-1985) could not be assigned to the two most frequently found trend segments. In total 11 models could be assigned to the two most common trend segments. Columns 5 and 6 show the direction of the jumps and the CRs of the trend segments (<1.0 or >1.0). Columns 7 and 8 characterize the number of joinpoints and the approximate duration for the respective trend segments. Columns 9 and 10 contain the results of the test for significance of the CRs (criterion 3) and for beta errors (criterion 4). Column 11 provides the overall assessment.

**Table 7.** Examination of the criteria based on the results of the models with 7 joinpoints.

| Year pair | Number of groups | Trend segments: period | Trend segments: number per period | Jump | CRs of the trend segments | Models with number JP | Average length of trend segments (in years) | Significance alpha | Significance beta | Overall assessment |
| --- | --- | --- | --- | --- | --- | --- | --- | --- | --- | --- |
| 1 | 2 | 3 | 4 | 5 | 6 | 7 | 8 | 9 | 10 | 11 |
| 1977/78 | 4 | 1952 to 1981-85 | 15 | ↘ | 15 out of 15: <1.0 | 2-5 | 30 | n.s.: 12 out of 15; p < 0.05: 3 out of 15 | .05 < p < 0.20: 6 out of 15 | No break |
|  |  | 1975-77 to 1981-87 | 8 | ↗ | 6 out of 8: >1.0 | 4-7 | 10 | n.s.: 8 out of 8; | .05 < p < 0.20: 0 out of 8 |  |
|  |  |  | 1<sup>a)</sup> | . | . | . | . | . | . | . |
| 1979/80 | 2 | 1952 to 1982/85 | 5 | ↘ | 5 out of 5: <1.0 | 2-5 | 30 | n.s.: 3 out of 5; p < 0.05: 2 out of 5 | .05 < p < 0.20: 1 out of 5 | No break |
|  |  | 1975/76 to 1987/88 | 6 | ↗ | 6 out of 6: >1.0 | 4-7 | 10 | n.s.: 6 out of 6 | .05 < p < 0.20: 0 out of 6 |  |
|  |  |  | 1<sup>a)</sup> | . | . | . | . | . | . | . |
| 1990/91 | 2 | 1980-83 until 2006/07 | 6 | ↘ | 6 out of 6: <1.0 | 2-5 | 25 | n.s.: 1 out of 6; p < 0.05: 5 out of 6 | .05 < p < 0.20: 1 out of 6 | No break |
|  |  | 1988 to 1993/94 | 6 | ↗ | 4 out of 6: >1.0 | 4-7 | 6 | n.s.: 6 out of 6 | .05 < p < 0.20: 0 out of 6 |  |
| 1997/98 | 4 | 1980-85 to 2005/07 | 6 | ↗ | 6 out of 6: >1.0 | 2-5 | 25 | n.s.: 6 out of 6 | .05 < p < 0.20: 4 out of 6 | No break |
|  |  | 1993/94 to 2003-08 | 8 | ↘ | 7 out of 8: <1.0 | 4-7 | 15 | n.s.: 8 out of 8 | .05 < p < 0.20: 2 out of 8 |  |
|  |  |  | 10<sup>b)</sup> | . | . | . | . | . | . | . |

Source: own compilation
<sup>a)</sup> another trend segment that does not fit into the present results system
<sup>b)</sup> further trend segments that do not fit into the present results system

For all year pairs, it can be observed that the jumps for long time periods often point in the opposite direction compared to the jumps for short periods. With few exceptions, the CRs are generally not significant. In none of the year pairs are the CRs significant for all populations.

The results for men in eastern Germany in 1990/91 deserve a special consideration. In models with 2 to 5 joinpoints, the CR is less than 1, although the empirical values show an increase from 1990 to 1991. This increase temporarily interrupted the general trend of decline during phase II. This phenomenon, which also occurred among women in eastern Germany, but to a lesser extent, can be attributed to the major social changes in eastern Germany in 1989/90/91. In the models with the higher number of joinpoints (women 4-7 joinpoints; men 5-7 joinpoints), the CRs turn into value ranges greater than 1, with p-values outside the interval of 0.05 to .20. The hypothesis of no significant jump can therefore not be rejected.

In summary, based on the results of the step-by-step procedure, it can be stated that there is no evidence of a methodologically induced structural break for any of the analysed year pairs.

The quality of suicide-mortality data is often discussed. In general, official statistics are assumed to underreport cases (see, for example, the systematic review (Tøllefsen et al., 2012). This underreporting is unlikely to systematically influence the trends in suicide mortality.

The reliability of the data from the official statistics of the GDR is also subject to debate. The mandated secrecy of the official suicide statistics suggests that the GDR leadership regarded this data as very problematic and sensitive (Grashoff, 2023; von den Driesch, 2021). However, there is no evidence of any deliberate falsification of the data (von den Driesch, 2021). According to Grashoff, the continued secrecy surrounding suicide statistics indicates that the data were not falsified (Grashoff, 2023): falsified figures would not have required secrecy.

## 5. Summary

The empirical results show that suicide mortality was highly dynamic in both parts of Germany, with frequent fluctuations in rates of increase and decrease. Despite elaborate and complex analyses, no indications were found of methodologically induced structural breaks as a result of the ICD version changes or the transition of the GDR statistical system to that of the FRG. In particular, the time-series data show no concerning effects suggesting changes in coding guidelines or customs in the course of this transition.

The analyses indicate that the compiled long time series for eastern and western German women and men are consistent. Methodological artefacts should therefore not be assumed if trend changes or shifts are found in future analyses.

## Data Availability

The official German cause-of-death data (FRG/western Germany: 1952-2022, GDR/eastern Germany: 1980-2022) analysed in this study were provided by the the Federal Statistical Offices (Online database https://www.gbe-bund.de/gbe). The cause-of-death data for the German Democratic Republic (1952-1979) were taken from the GESIS database Suicide, demographic, socio-structural, infrastructural and crime statistics of the German Democratic Republic (https://data.gesis.org/sharing/#!Detail/10.7802/1.2010, https://doi.org/10.7802/1.2010)

https://data.gesis.org/sharing/#!Detail/10.7802/1.2010

## Footnotes

1 We use the following terms for the two parts of Germany in the respective time periods: 1952-2022: eastern Germany and western Germany; 1952-1990: GDR and FRG; 1991-2022: new federal states and old federal states

## Notes

### Competing Interest Statement

The authors have declared no competing interest.

### Funding Statement

This research was supported by the German Federal Ministry of Education and Research (Grant number: 01UJ1911CY). The funding body had no role in the design of the study and collection, analysis, and interpretation of data, in writing the manuscript, and in the decision to submit it for publication.

